# African-Ancestry Associated Gene Expression Signatures and Pathways in Triple Negative Breast Cancer, a Comparison across Women of African Descent

**DOI:** 10.1101/2022.01.27.22269747

**Authors:** Rachel Martini, Princesca Delpe, Timothy R. Chu, Kanika Arora, Brittany Lord, Akanksha Verma, Yalei Chen, Endale Gebregzabher, Joseph K. Oppong, Ernest K. Adjei, Aisha Jibril, Baffour Awuah, Mahteme Bekele, Engida Abebe, Ishmael Kyei, Frances S. Aitpillah, Michael O. Adinku, Kwasi Ankomah, Ernest B. Osei-Bonsu, Dhananjay Chitale, Jessica M. Bensenhaver, Saul David Nathanson, LaToya Jackson, Evelyn Jiagge, Lindsay F. Petersen, Erica Proctor, Kofi K. Gyan, Lee Gibbs, Zarko Monojlovic, Rick Kittles, Jason White, Clayton Yates, Upender Manne, Kevin Gardner, Nigel Mongan, Esther Cheng, Paula Ginter, Syed Hoda, Olivier Elemento, Nicolas Robine, Andrea Sboner, John Carpten, Lisa Newman, Melissa B. Davis

**Affiliations:** Department of Surgery, Weill Cornell Medical College, New York City, NY, USA; (RM), (BL), (KG), (LN), (MD); Department of Genetics, University of Georgia, Athens, GA, USA; Englander Institute for Precision Medicine, Weill Cornell Medical College, New York City, NY, USA; (PD), (AV), (OE); New York Genome Center, New York City, NY, USA; (TC), (KA), (NR); Department of Public Health Sciences, Henry Ford Health System, Detroit, MI, USA; (YC), (LJ), (EJ); Department of Biochemistry, St Paul’s Hospital Millennium Medical College, Addis Ababa, Ethiopia; (EG); Department of Surgery, Komfo Anokye Teaching Hospital, Kumasi, Ghana; (JO), (FA); Department of Pathology, Komfo Anokye Teaching Hospital, Kumasi, Ghana; (EA); Department of Pathology, St. Paul’s Hospital Millennium Medical College, Addis Ababa, Ethiopia; (AJ); Directorate of Oncology, Komfo Anokye Teaching Hospital, Kumasi, Ghana; (BA), (EO); Department of Surgery, St. Paul’s Hospital Millennium Medical College, Addis Ababa, Ethiopia; (MB), (EA); Department of Surgery, Kwame Nkrumah University of Science and Technology, Kumasi, Ghana; (IK), (MA); Directorate of Radiology, Komfo Anokye Teaching Hospital, Kumasi, Ghana; (KA); Department of Pathology, Henry Ford Health System, Detroit, MI, USA; (DC); Department of Surgery, Henry Ford Health System, Detroit, MI, USA; (JB), (SN), (LP), (EP); Department of Translational Genomics, Keck School of Medicine, University of Southern California, Los Angeles, CA, USA; (LG), (ZM), (JC); Department of Population Sciences, City of Hope, Duarte, CA, USA; (RK); Department of Biology, Tuskegee University, AL, USA; (JW); Center for Cancer Research, Tuskegee University, AL, USA; (CY); Department of Pathology, University of Alabama at Birmingham, Birmingham, AL, USA; (UM); O’Neal Comprehensive Cancer Center, University of Alabama at Birmingham, Birmingham, AL, USA; Department of Pathology and Cell Biology, Columbia University, New York, NY, USA; (KG); Biodiscovery Institute, University of Nottingham, Nottingham, UK; (NM); Department of Pharmacology, Weill Cornell Medical College, New York City, NY, USA; Department of Pathology and Laboratory Medicine, Weill Cornell Medical College, New York City, NY, USA; (EC), (PG), (SH), (AS); Institute of Computational Biomedicine, Weill Cornell Medical College, New York City, NY, USA

**Keywords:** African ancestry, health disparities, triple negative breast cancer, gene expression, genetic ancestry

## Abstract

Women of sub-Saharan African ancestry have disproportionately higher incidence of aggressive, early-onset Triple Negative Breast Cancer (TNBC), and TNBC mortality across all race groups. Population-based comparative studies show racial differences in TNBC tumor biology, with higher prevalence of basal-like and Quadruple-Negative subtypes in African Americans (AA). However, most investigations relied on self-reported race (SRR) of primarily United States (US) populations. However, given that genetic admixture in AAs is extremely heterogenous, and race-correlated social determinants can translate into biological differences, the true association of African ancestry with TNBC biology and gene expression is currently unclear. To address this, we conducted RNAseq on an international cohort of AAs, west and east Africans with TNBC. Using genetic ancestry estimation in this African-enriched cohort, we identified 613 genes associated with African ancestry and more than 2200 genes associated with regional-level African ancestry. Functional enrichment and deconvolution revealed tumor-associated immune cell infiltration and activity.

**STATEMENT OF SIGNIFICANCE:** Using a rigorous ancestry quantification process, we show that TNBC has ancestry-associated gene expression profiles, linked to immunological landscapes, which may contribute to racial differences in clinical outcomes. This is the first study to show the definitive link to tumor immunological landscape, associated with African ancestry, using a multiethnic African-enriched cohort.

## INTRODUCTION

Breast cancer (BC) is the most frequently diagnosed cancer among women globally, and the leading cause of cancer-related death among women^1,2^. Despite having lower BC incidence, mortality rates are among the highest across most sub-Saharan African nations, compared to other nations worldwide. While poorer survival is typically attributed to advanced-stage disease at presentation and limited access to treatment options in Lower-Middle Income Countries (LMIC)^2^, triple negative BC (TNBC) incidence rates across African nations represent approximately 33% of BC diagnoses compared to less than 20% in other nations^3,4^ with highest incidence of TNBC in west African nations compared to east African nations^3,5,6^. Globally, overall BC mortality and TNBC burden appears higher across the African diaspora at-large, corresponding with a higher prevalence of TNBC disease among women with African ancestry^6^, who reside in nations throughout Europe^7,8^, South Africa, and admixed African American populations in the US^5,9,10^. We previously reported a higher risk of TNBC, compared to other types of breast cancer, associated with west African ancestry^5,6^. Therefore, we hypothesized that there may be genetic drivers associated with west African ancestry that predispose and/or lead to aggressive breast cancer, including TNBC.

TNBC continues to have the worst prognosis of BC subtypes, and the worst survival outcomes due to lack of targeted therapy options for these tumors^11,12^. Given TNBC incidence rates across the African diaspora, our efforts in oncologic anthropology have shifted to a molecular focus to uncover and characterize the influence of African ancestry on BC disease etiology and progression^4,13,14^. Previous comparative BC studies among patients of diverse race groups have focused on comparing tumors from African American (AA) and European American (EA) self-reported race (SRR) groups in the US. While there were inherent limitations of cohort size and heterogeneity of race, these approaches were useful in determining that broad biological differences do exist across diverse patient populations^15,16^. Some of these discoveries included race-group distinctions in genomic differences in frequencies of Single Nucleotide Variants (SNVs)^6,17,18^, somatic tumor mutation signatures^19,20^, structural copy number variations (CNVs)^21^ and differences in DNA methylation patterns in both ER+ and ER-tumors^22^. Our work and others has uncovered racial differences in gene expression that revealed distinctions in immune response signatures, repeatedly across independent cohorts, implicating differences in the Tumor Microenvironment (TME)^16,23^ as a possible cause of outcome disparities. The emerging promise of curative immunotherapies in overcoming treatment resistance in TNBC highlights an important opportunity to target the immune microenvironment and increasing relevance to racial group differences to overcome disparities^24,25^. However, there are limitations to using self-reported race in genomic studies, mainly due to complexity in genomic backgrounds of admixed groups.

Our recent work was the first to use quantified genetic ancestry in admixed AA women to identify African ancestry-specific gene expression differences in TNBC tumors compared to EA women, which we also showed overlapped with SRR-associated gene networks^26^. Of the African ancestry-associated genes, 48.1% were distinct from the SRR-associated genes, indicating the functional influence of the genetic ancestry background upon gene expression, apart from SRR alone. Similarly, a recent study from our collaborators characterized gene expression of TNBCs from Bantu tribe from Kenya and found Bantu population-specific gene expression signatures as compared to TNBCs of AA and EA TNBCs^14^. However, the implications of ancestry are still untested, lacking the inclusion of the contemporary and appropriate representative ancestry groups that are specific and relevant to the admixed patient groups.

Therefore, our current study utilized an African-enriched international cohort from the International Center for the Study of Breast Cancer Subtypes (ICSBCS)^27^, which will help to resolve a more precise understanding of genetic influences associated with African ancestry in race-associated gene signatures. We have measured the influence of African ancestry on TNBC tumor biology, derived from gene expression differences, that includes west African/Ghanaian and east African/Ethiopian women with TNBC, compared to admixed AAs. Our rationale was based on firmly in prior studies indicating that shared African ancestry harbors both unique genetic risk of TNBC tumor etiology and the distinct gene signatures of TNBC among women of the African diaspora^6,18,26^. We identified both African ancestry-associated gene expression signatures, and TME cell type differences from bulk RNA sequence (RNAseq) data. We demonstrate that inclusion of native Africans with admixed AA patients, who share the same genetic ancestry, can overcome population complexity to help deduce the shared genetic drivers observed in race-group differences and discern these from environmental or other exogenous drivers of gene expression changes. We have identified subpopulation differences in gene expression between east vs west African ancestry lineages, which can be applied throughout population studies of the African diaspora in Europe^7,8^, the US^5,9,10^ and abroad (i.e. Afro-Latinx and Afro-Caribbean).

## RESULTS

### Characterization of ancestry profiles reveals complex admixture in African and AA cohort

We estimated the global genomic ancestry for each patient in our cohort to determine the varying levels of admixture, based on 1000 Genomes superpopulation and subpopulations (**Supplemental Table S1**)^28^. Our cross-sectional set of 148 TNBC cases includes 66 AAs and 41 EAs, enriched with 13 west African Ghanaians, and 22 east African Ethiopians, with four individuals who declined to report SRR (**Supplemental Figure S1**). African ancestry comparisons indicated significant differences in African ancestry across our cohort (ANOVA *p* < 0.001), with Ghanaian patients having the highest levels of African ancestry (median 97.3%), and AA having an average 15% less African ancestry (median 82.6%). Ethiopian patients had a surprisingly lower African ancestry (median 43.0%), with nearly equal amounts of European ancestry (median 43.5%), which is consistent with previous anthropological studies^29–31^. (**Figure 1A, Supplemental Table S2**). Our EA patients generally show exceptionally low levels of African ancestry (median 2.4%); however, three self-identified EA patients had between 30-80% African ancestry.

**Figure 1.**
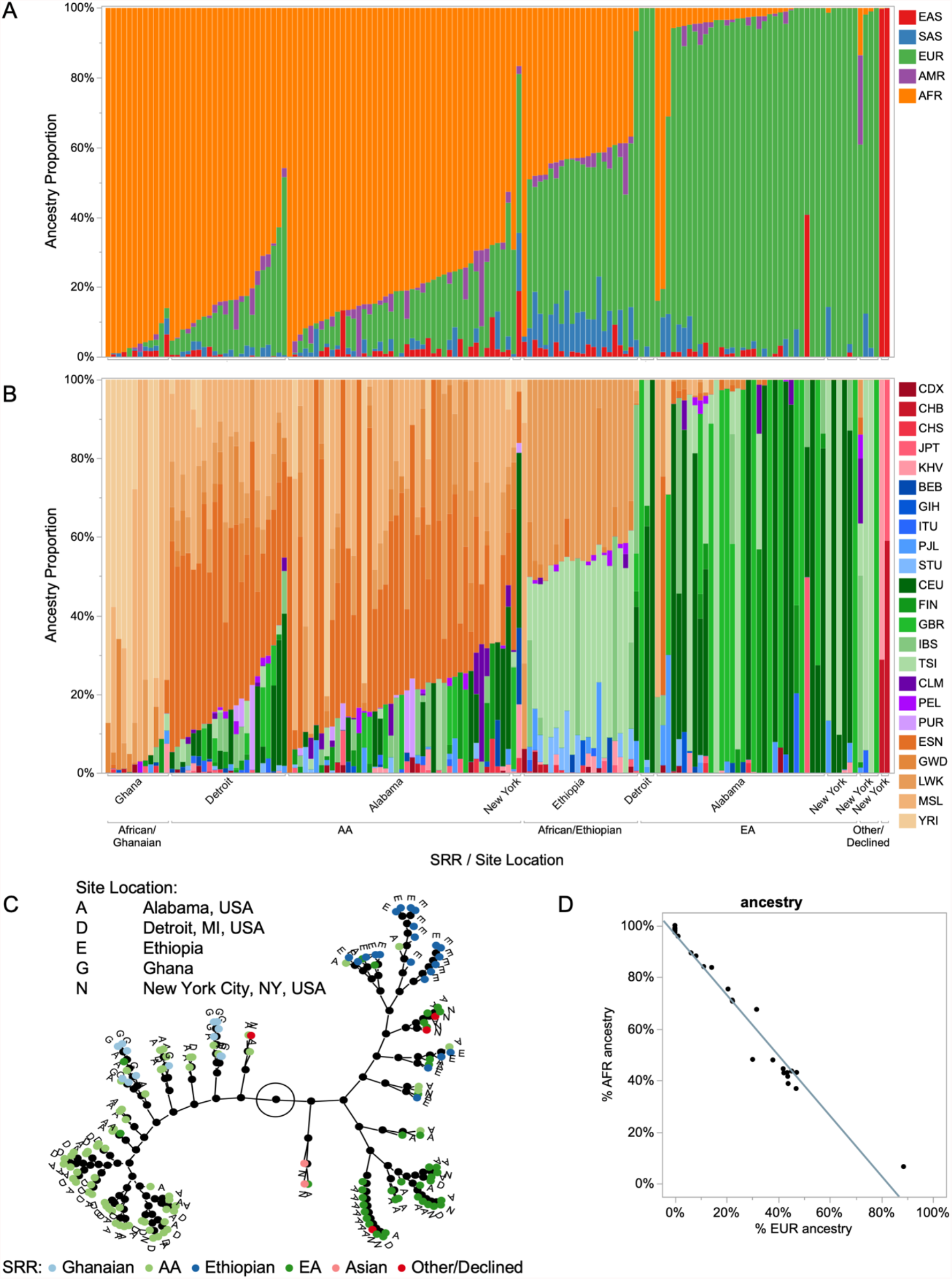
Estimated genetic ancestry distribution in an African-enriched TNBC RNAseq cohort. Genetic ancestry was estimated from genotypes of the ancestry-informed markers, obtained from our RNAseq alignments, where we have (A) superpopulation ancestry estimations, relative to the 1000 Genomes superpopulation populations, and (B) subpopulation ancestry estimations for each individual in our cohort. In both (A) and (B), each column represents an individual in the cohort, where estimated ancestry from a given superpopulation or subpopulation is shown on the y-axis, and the x-axis is annotated by SRR and location. Superpopulation populations in (A) are East Asian (EAS, red), South Asian (SAS, blue), European (EUR, green), American (AMR, purple) and African (AFR, orange). Subpopulations in (B) are shown in variations of their corresponding superpopulation population color (i.e. African populations are in varying shades of orange). Samples are ordered by decreasing AFR ancestry (x-axis left to right: African/Ghanaian (Ghana), AA (Alabama, Detroit, New York), African/Ethiopian (Ethiopia), EA (Alabama, Detroit, New York), Other/Declined (New York) and Asian (New York). (C) Constellation plot showing phylogeny of samples based on ancestry estimations. SRR of samples are indicated by the colored dots (Ghanaian = light blue, AA = light green, Ethiopian = dark blue, EA = dark green, Asian = light pink, Other/Declined = dark pink). Site location of samples are annotated next to the color dots (A = Alabama, USA, D = Detroit, MI, USA, E = Ethiopia, G = Ghana, N = New York City, NY, USA). (D) Scatterplot showing inverse correlation of AFR and EUR ancestry in our gene expression cohort.

For more precise ancestry estimations that reflect regional origins, we estimated genetic ancestry for five African subpopulations, which includes four populations representing west African Ancestry, including: Esan in Nigeria (ESN), Yoruba in Ibadan, Nigeria (YRI), Gambian in Western Divisions in the Gambia (GWD) and Mende in Sierra Leone (MSL). There was only one population representing east Africa in 1000 Genomes, Luhya, in Webuye, Kenya (LWK). (**Figure 1B, Supplemental Table S1**). As anticipated, AA patients presented with African ancestry primarily of west African origin, which included ESN (median 36.1%) and MSL (median 19.7%) ancestry, with less than 10% estimated east African ancestry (LWK median 7.5%). Interestingly, the heterogeneity of African origin within AAs is more extensive and wide-ranging than the origin of African ancestry in Ghanaians or Ethiopians, where the amount of specific subpopulation ancestry can range from 0% to 90% for a given individual, indicating the complex diversity of African admixture in AAs. African patients’ subpopulation ancestry is highly correlated with their regions of origin, where Ghanaian ancestry is overwhelmingly correlated with the west African reference groups from YRI (median 66.0%), and MSL (median 24.1%) and Ethiopian patients have almost exclusively east African ancestry, represented as LWK (median 43.0%). Phylogeny of patients based on estimated genetic ancestry shows separation of Ghanaian and AA patients from Ethiopian and EA patients (**Figure 1C**). Interestingly, the European ancestry in our Ethiopian patients was primarily Italian (Toscani in Italia (TSI), median 41.2%). Ethiopian patients also showed substantial levels of East and South Asian ancestry (EAS median 1.9%, SAS median 9.0%), with more SAS compared to other SRR groups. All of these admixture revelations are consistent with the social histories of each SRR group and reflect the diversity across the African diaspora^29–33^.

### Influence of ancestry in gene expression profiles of TNBC tumors results in ancestry-associated differential immune signatures

To investigate African ancestry-specific gene expression profiles in our ICSBCS TNBC samples, we isolated our analyses to patients with significant (>35%) AFR ancestry. As previously described^26^, we performed gene-by-gene linear regression, using genetic ancestry as a continuous variable, which determined ancestry-associated gene expression. We identified gene signatures associated with AFR (n = 613) and EUR (n = 345) ancestry (*p* < 0.001), with 293 genes shared between these gene signatures (**Figure 2A**) (**Supplemental Table S3**). Given the significant inversely correlated AFR vs EUR ancestry in our patient cohort (**Figure 1D**), we compared the polarity of gene expression levels of the 293 overlapping genes and found that genes upregulated in association with AFR ancestry are conversely downregulated in association with EUR ancestry (**Figure 2B**). This may represent genes that have expression drivers that are ancestral informative variants, isolated to certain ancestry groups.

**Figure 2.**
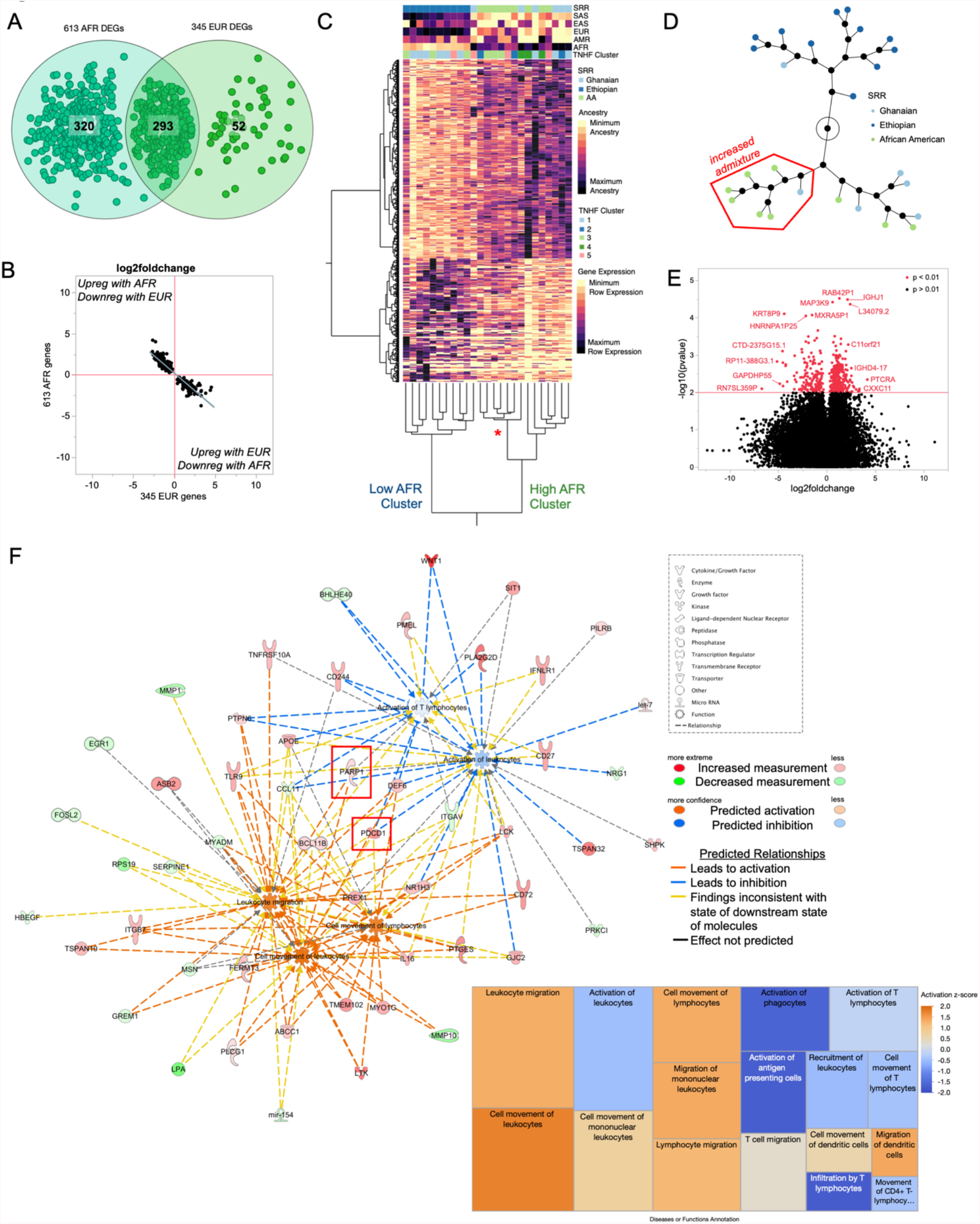
African ancestry associated genes show enrichment in immune response. (A) Venn diagram of ancestry-associated genes identified from AFR and EUR genetic ancestry logistic regression model, where ancestry was used as a continuous variable. (B) Scatterplot showing log2foldchange of 293 overlapping genes from AFR- and EUR-associated gene signatures. The upper left quadrant represents those genes upregulated with increasing AFR ancestry (positive log2foldchange on y-axis), and subsequently downregulated with increasing EUR ancestry (negative log2foldchange on x-axis). (C) Unsupervised hierarchical clustering of 613 AFR QGA associated DEGs. Columns represent individuals, where SRR, QGA estimates and TNBC subtypes are indicated in the colormap at the top of the heatmap. Rows represent DEGs, where lighter yellow indicated minimum row expression, and darker purple shows maximum row expression. (D) Constellation plot representing nodal structure of individuals from (C), where points are colored by SRR (Ghanaian = light blue, Ethiopian = dark blue, AA = light green). Node highlighted by red box indicated increased admixture node, highlighted in (C) by the red star. (E) Volcano plot of AFR-associated genes, where 613 significant genes area shown in red. (F) Network and treemap diagram of Ingenuity Pathway Analysis (IPA) immune cell trafficking disease and function terms, where the 613 AFR gene signature was enriched (p value range of terms = 0.0119 – 0.000502). Genes in red or green are upregulated or downregulated among High AFR individuals, respectively. Genes and treemap boxes in orange represent a positive z-score (predicted activation), and those in blue represent a negative z-score (predicted inhibition).

Unsupervised hierarchical clustering of the 613 AFR-associated genes separated patients into two distinct clusters, correlating with levels of AFR ancestry, which we denote as a Low AFR cluster, including primarily Ethiopian TNBC patients, and a High AFR cluster, including primarily AA and Ghanaian patients (**Figure 2C**). The High AFR subcluster includes two sub nodes representing ESN, MSL and LWK ancestry, (6/9 AA and 1/6 Ghanaians), and a second node representing YRI, GWD and ESN ancestry, (3/9 AA and 4/6 Ghanaians) (**Figure 2C, red asterisk; Figure 2D, red box**). The sub nodes reflect differences in origin of AFR ancestry composition observed among west Africans and AAs in our cohort.

We calculated AFR-associated DEGs (**Figure 2D**) for functional pathway enrichment, using the log2fold change between the distinct High AFR and Low AFR clusters to measure differential expression. Top canonical pathways included some previously implicated processes in race-group comparisons; such as: RNA post-transcriptional modification through spliceosomal cycle pathway enrichment (*p* value = 0.0002, z-score = 3.804)^34^, cell to cell and extracellular matrix interactions in the integrin signaling pathway (*p* value = 0.004, z-score = 0)^35^ and chronic inflammation in atherosclerosis signaling (*p* value = 0.006, no z-score predicted)^36^. Upregulation of WNT family member genes drive enrichment in a colorectal cancer metastasis signaling pathway (*p* value = 0.004, z-score = −0.302)^37^, and HOTAIR regulatory pathway (*p* value = 0.006, z-score = −0.707). One of the top enriched functions identified was Immune Cell Trafficking (*p* value range of sub-terms 0.0119 – 0.000502) (**Figure 2F**). Specifically, there was a predicted increase in signals relating to immune cell movement and migration, but conversely a predicted inhibition of signals relating to immune cell activation. This finding was of particular interest, given our previous findings related to DARC-regulated immune cell infiltration, associated with race-groups^38^.

### Resolution of subpopulation African ancestry influence on gene expression signatures

We determined a higher resolution of African subpopulation origins to harness the shared genetic diversity and identify subpopulation associated gene signatures. Specifically, we utilized 1K Genomes African subpopulation ancestry (**Figure 1**) estimates for west African (YRI, ESN, GWD and MSL), and east African (LWK) populations (**Supplemental Table S1**). By repeating the gene-by-gene statistical model with African subpopulations, we identified a combined 2567 genes associated with the five African population groups (**Figure 3A**). African subpopulation-specific gene associations included 338 YRI genes, 643 ESN genes, 201 GWD genes, 146 MSL genes, and 1229 LWK genes (**Supplemental Table ST4**). These gene lists included, but extended beyond, the genes identified in the AFR superpopulation ancestry analysis (**Figure 3A)**. Surprisingly, there were no DEGs shared among all five populations, suggesting there are unique gene expression drivers from each ancestry group. Though, a small fraction of each individual west African subpopulation genes were shared with the east African LWK population (total n = 210). As what may be anticipated, we found the largest overlap of African subpopulation-associated genes were shared between ancestry groups that are geographically adjacent nations (29.0% of YRI and 48.8% of GWD shared between these populations). However, the closest west African groups, YRI and ESN of Nigeria, did not share any associated genes.

**Figure 3.**
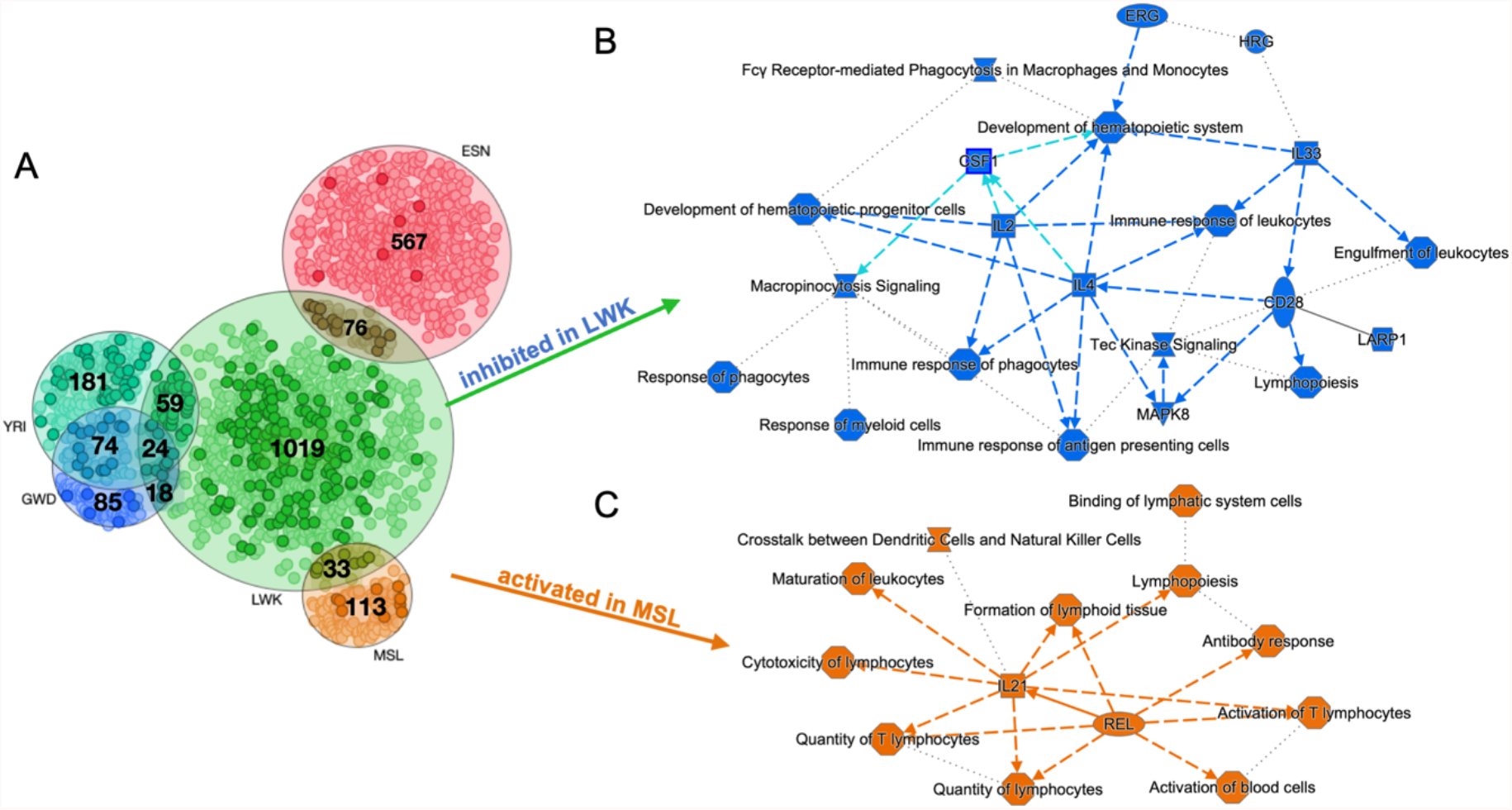
African subpopulation associated genes are also enriched in immune response. (A) Venn diagram of unique and overlapping gene signatures associated with LWK, ESN, MSL, YRI and GWD ancestry, respectively. Dots that are bolded are genes that overlap with the 613 AFR-associated gene signature. IPA analysis of (B) LWK-associated and (C) MSL-associated genes. (B) Colors in blue indicate inhibition of regulators, disease/function terms and canonicals pathways among individuals with increasing LWK ancestry. (C) Colors in orange indicated activation or regulators, disease/function terms and canonical pathways among individuals with increasing MSL ancestry.

Therefore, we considered which SRR/nationality groups carried the specific subpopulation ancestry and therefore which groups these gene signatures may be found. The east African population, LWK with the largest set of associated genes (n = 1229) predominantly represented our Ethiopian patients with a small portion AA ancestry represented by LWK (median ~8%). Pathway analysis predicted decrease in immune response related function in east African ancestry gene sets, including inhibition of CSF-1 and various interleukins, CD28 and lymphopoiesis, and the canonical tec kinase signaling pathway (**Figure 3B**). This inhibitive effect of LWK ancestry on these functions clearly distinguishes the differences in tumor biology between west vs east Africans and suggests important differences in immune cell development, response, and activation. MSL ancestry was predominantly found among AAs and Ghanaians, and included genes (n=146) that also involved function of immune response signals that suggest activation of immune-related functions (**Figure 3C**). Specifically, there was activation of REL and IL21, both playing important roles in immune response regulation. To validate our findings, we conducted a re-analysis of the AA subset of our previously published cohort, and identified AFR subpopulation-associated gene signatures enriched for pathways that involve immune function (**Supplemental Figure S2**).

### Expressed signatures of immune cell enrichment are associated with African ancestry

We estimated immune cell populations and overall tumor-associated leukocyte (TAL) abundance with deconvolution and cell-type enrichment methods, CIBERSORTx^39^ and xCell^40^. Absolute scores, which are the sum of all estimated immune populations, was significantly higher among patients with high AFR ancestry, compared to low AFR patients (**Figure 4A**, *p* = 0.0076). Specific immune cell populations driving the bulk of these differences included naïve B cells, CD8+ T cells, helper T cells, regulatory T cells, and activated mast cells (**Figure 4B**). Ancestry-specific association testing show that these same immune cell populations are significantly associated with African ancestry (**Figure 4C**). The African ancestry pathway enrichment had indicated stimulation of ‘migration/movement’ functions with repression of ‘cell-type activation’ functions associated with African ancestry, and interestingly, the largest contributing cell type the AFR-associated TALs are naïve B cells, a non-activated immune cell population. Conversely, an activated immune cell population, activated mast cells, are more prominent in tumors of patients with low AFR ancestry. In an independent cell-type enrichment analysis using xCell^36^, we replicated the CIBERSORTx findings of AFR-associated immune cell infiltration, and further discerned that the specific T populations associated with African ancestry are CD8+ T cells, and CD8+ T effector memory cells (**Supplemental Figure S3**, *p* < 0.05), which is concordant with the association of CD8+ T cells from CIBERSORTx deconvolution (**Figure 4C**, *p* < 0.05).

**Figure 4.**
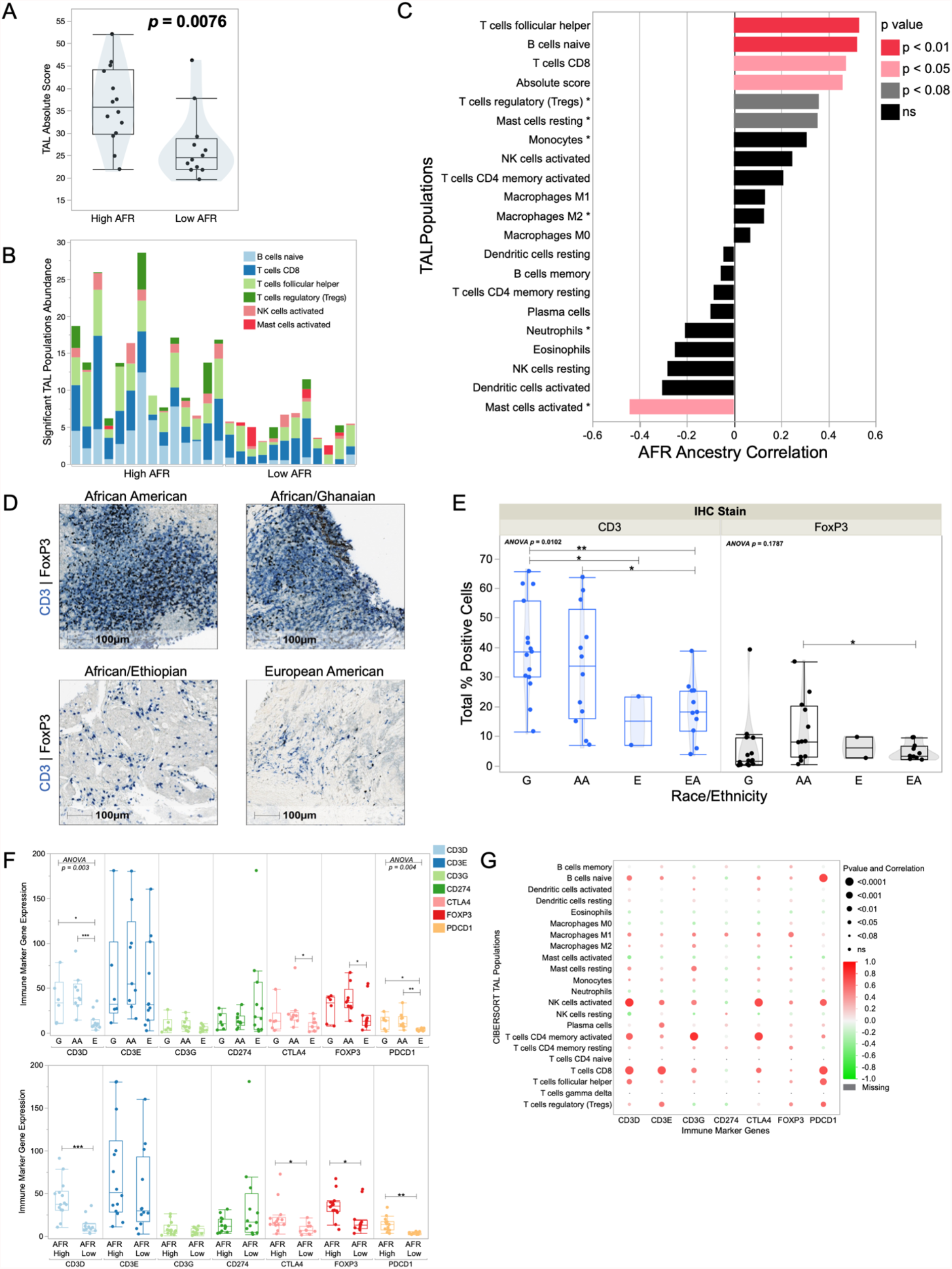
Immune deconvolution of bulk tumors shows enrichment of immune cells among High AFR ancestry tumors. (A) Box plot of TAL absolute score among High AFR and Low AFR samples (student’s t-test *p* = 0.0076). (B) Stacked bar chart of TAL populations significantly different between AFR High and AFR Low samples. (C) Correlation of African ancestry and CIBERSORTx tumor-associated leukocyte (TAL) populations. Significant correlations are highlighted in shades of red. TAL populations with a star represent immunosuppressive cell populations^41^. (D) Representative IHC images of CD3 (blue) and FOXP3 (black) staining in African American (upper left), Ghanaian (upper right), Ethiopian (lower left) and European American (lower right) TNBC cases. € Boxplots of percent positive CD3 (blue) and FOXP3 (black) stained cells from IHC images by SRR groups. ANOVA p values and paired student’s t-tests (**= *p* < 0.01, * = *p* < 0.05) are reported on the plot. (F) Box plot of gene expression of CD3D (light blue), CD3E (dark blue), CD3G (light green), CD274 (dark green), CTLA4 (light pink), FOXP3 (dark red) and PDCD1 (light orange), across SRR groups (G = Ghanaian, AA = African American, E = Ethiopian) or AFR cluster groups (AFR High, AFR low). Significant ANOVA p values and paired student’s t-tests (*** = *p* < 0.001, ** = *p* < 0.01, * = *p* < 0.05) are reported. (G) Correlation of immune marker gene expression (bottom) and CIBERSORTx tumor associated leukocyte populations (left). Positive correlation is shown in red, and negative correlation is shown in green. Size of the dot represents the significance of the correlations.

To validate the RNAseq-based immune cell estimations, we used clinical-grade IHC marker assays to score the infiltration of immune cells in an independent set of ICSBCS TNBC patients (n=40), distributed across each ethnicity group represented in the RNAseq cohort. We found similar immune cell infiltration trends across race/ethnic groups, with Ghanaian and AA tumors having higher counts of CD3+ and FOXP3+ cells, compared to Ethiopian and EA tumors (**Figure 4D**). CD3+ cells showed significant variation across all race/ethnic groups (ANOVA *p* = 0.0102, **Figure 4E**), with significant pair-wise differences between Ghanaians and Ethiopians (*p* = 0.0457) and between AA and EA (*p* = 0.0379). We then verified that the IHC T cell markers’ corresponding gene (*CD3D* and *FOXP3*) RNA expression matched IHC findings among SRR groups (**Figure 4F**) and found that the greatest differences were found between AFR ancestry, overshadowing the SRR group differences (**Figure 4F**). This indicates that the genetic ancestry is the driver of these immunological differences, not SRR.

FOXP3 expression in T-regs is correlated with a suppressive immune tumor microenvironment (TME), suggesting patients with higher African ancestry may have TME that is more suppressive vs stimulating, compared to patients of European ancestry. Therefore, we investigated immune suppressive vs stimulating TME marker associations^41^ with African ancestry by comparing the relative expression of several well-known immune checkpoint genes, including, *CD274* (PD-L1 marker), *CTLA4*, and *PDCD1* (PD-1 marker) (**Figure 4F**). We found that *PDCD1* was significantly associated with AFR ancestry and SRR (ANOVA *p* < 0.01), with both Ghanaian (mean 12.42) and AA (mean 13.72) tumors patients having 4x higher expression than Ethiopian patients (mean 3.68). This further suggests an immune suppressive tumor environment being associated specifically with west African ancestry, as opposed to east African or European ancestry. To ensure these immunosuppressive markers’ gene expression patterns were derived from the immune cell population within the bulk tumor, we tested the correlation of specific immune cell estimates from CIBERSORTx with the CTLA4, CD3D and PDCD1 markers. We found each correlated with the abundance of relevant T-cell subtypes, indicating these cells are the likely source of the RNA expression (**Figure 4G**).

### TNBC subtyping reveals ancestry bias in composition of mosaic heterogeneity

TNBC tumors can be categorized into subtypes that have previously been shown to predict distinct clinical, as established outcomes^42,43^. The initial report of these subtypes by the landmark study introducing the well-known Vanderbilt TNBCtype tool. At its inception, the tool designated tumors into five distinct subtypes, correlated to the gene expression signatures of a tumor training set: including, basal-like 1 (BL1), basal-like 2 (BL2), luminal androgen receptor (LAR), mesenchymal (M), mesenchymal-stem like (MSL) and immunomodulary (IM). An additional ‘subtype’ category harbors tumors with ‘unsure calls’ (UNS), which describes a tumor with either multiple subtype correlations, or no positive correlations with any of the established phenotypes. Further consideration of the histological context of these tumor subtypes determined that the MSL and IM subtypes are stromal and tumor-associated immune derived, rather than distinct phenotypes. The correction currently applied to IM or MSL tumors are to manually re-assign the calls with their second-most significant correlated call^43^. Distribution of the original Vanderbilt TNBC subtypes in our cohort and found tumors from Ghanaian and Ethiopian patients were more often BL1, and tumors from AA patients were more often IM subtype (**Figure 5A, top)**. Interestingly, all IM tumors were from the High AFR ancestry Ghanaian and AA individuals, indicating the strong tumor immune signatures in these ancestry groups. After the suggested re-assignment of the IM and MSL subtypes, AAs had a predominance of UNS calls, indicating an unresolved heterogeneity that would not allow designation of a single subtype (**Figure 5A, middle**). Therefore, to ascribe a biological phenotype to these tumors, we employed a previously described median-ranking^26^ which excludes the confounding influence of the immune signature genes. Our results indicate that BL1 is the predominant subtype for Ghanaians, and M is the predominant subtype for both AA and Ethiopians (**Figure 5A, bottom**).

**Figure 5.**
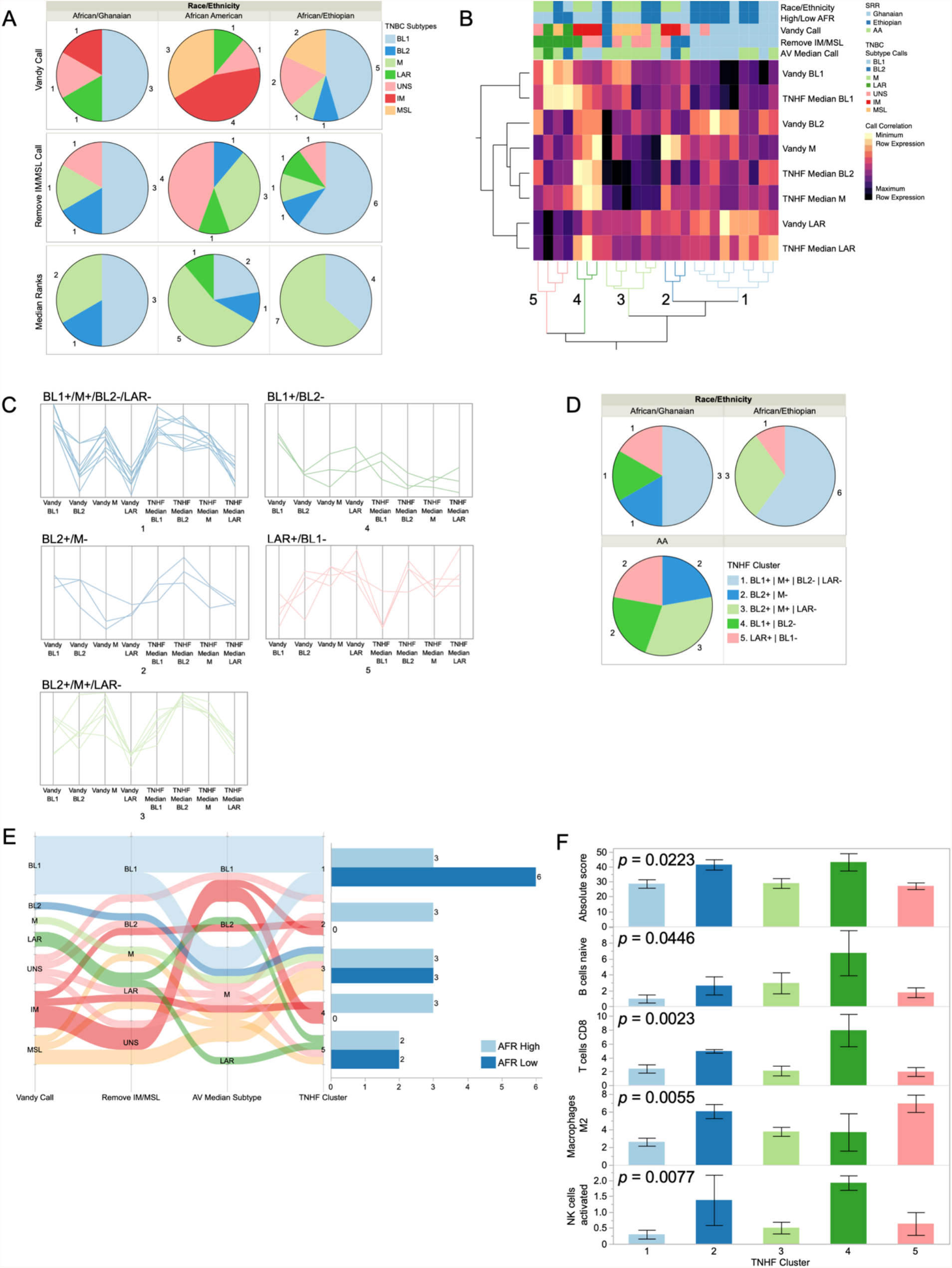
TNBC subtyping reveals heterogeneity of tumors. (A) Pie charts showing distribution of TNBC subtypes across SRR groups for the TNBCtype initial call (Vandy Call, top row), TNBCtype call after removing/re-assigning IM and MSL calls (middle row), and the calls using our median ranks method (bottom row). (B) Heatmap of correlations from the Vanderbilt TNBC subtyping tool, and our median ranks calling for TNBC subtypes. Color map at the top indicates self-reported race/ethnicity, High or Low AFR cluster sample, Vanderbilt TNBC subtyping call, Vanderbilt call after removal of IM/MSL, and our median ranks call. Samples clustered into 5 groups, which are color-coded and labelled 1-5 on the dendrogram at the bottom. (C) Line plot depicting positive and negative correlations with the Vandy tool and the median ranking subype calls in each of the TNHF clusters. (D) Pie charts showing distribution of TNHF clusters across SRR groups. (E) Sankey plot showing distribution of calls from initial Vanderbilt TNBCtype results to Vanderbilt call after removal of IM/MSL, to our Median Ranks method, to the final TNHF clusters from (B). Color coding is based of initial Vandy Call (left). Bar chart to the right shows the number of tumors from AFR High or AFR low in a given cluster. (F) Stacked bar chart of CIBERSORTx TAL populations in each of the TNHF clusters.

Given the heterogeneity of TNBC tumors among AA patients, we also employed our previously established Triple-Negative Hetero Fluid (TNHF) subtyping method^26^, which allows for assignment of multiple subtypes in a tumor’s composition. Our method also considers the exclusion of certain subtypes to establish unique combinations of subtype composition. Unsupervised hierarchical clustering correlated the various rankings of subtype categories among all these tools for each tumor and resulted in distinct clusters of similar subtypes based on the heterogenic designations, resolving into five distinct nodes (**Figure 5B and 5C**). Cluster 1 is the largest node composed of BL1+/M+/BL2−/LAR-tumors from Ghanaians and Ethiopians. Cluster 2 represents AFR high cases only and are BL2+/M−. Cluster 3 tumors are BL2+ and M+ tumors originating from AA and Ethiopian tumors. Cluster 4 are also AFR high cases only, and BL1+/BL2−. Lastly, cluster 5 includes LAR+ tumors derived from each of the patient groups (**Figure 5C-E**). Further evaluation of heterogeneous clusters 2 and 4 show all samples were initially called IM (with 1 initially called UNS), and all belong to the high AFR cluster (**Figure 5E**). Correlation of TAL populations with TNHF cluster 2 and 4 designations show enrichment in immune cell populations (**Figure 5F**), including B cells, CD8+ Tcell, M2 macrophages and NK cells.

### Genes associated with self-reported race, are involved in comorbidity pathways

In the interest of determining any distinct impact of racial social constructs on the biological phenotypes of tumors, we also investigated the functional pathway enrichment of SRR-associated genes. We hypothesized that pathways associated with SRR-associated gene signatures would yield different findings from our ancestry-associated gene signatures. The SRR-associated gene signature identified 1071 differentially expressed across SRR groups, and these were compared to our 613 AFR-associated genes and 345 EUR-associated genes to determine any overlaps in the gene signatures (**Figure 6A**). The overlap of ancestry-associated genes (AFR and/or EUR) with SRR genes included 320 genes, where 751 genes were uniquely associated with SRR. These differences indicate the importance of both an individual’s genetic ancestry and SRR on gene expression signatures, even in the context of tumor gene expression profiles.

**Figure 6.**
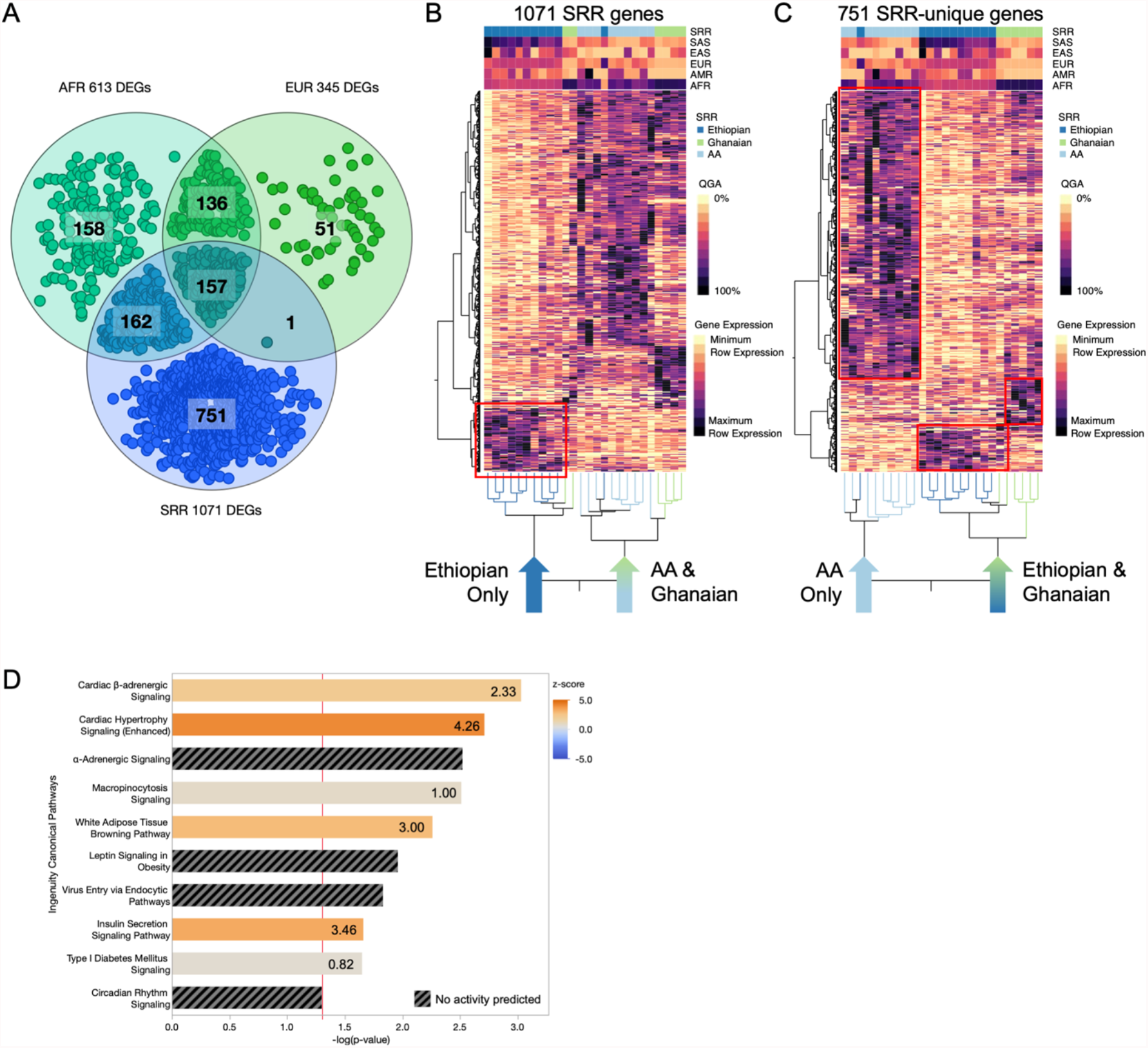
SRR-unique gene signature enriched in co-morbid canonical pathways. (A) Venn diagram depicting overlap of AFR-, EUR- and SRR-associated genes. (B) Unsupervised hierarchical clustering of the 1071 SRR-associated genes. (C) Unsupervised clustering of 751 genes unique to SRR. In both (B) and (C), columns represent individuals, where SRR and QGA is showing in the color map at the top, and rows represent DEGs. Node structure of individuals is shown at the bottom of the heatmaps, where clustering was individual node structure significantly changed between panels (B) and (C). (D) Comparing gene expression values from panel (C) node structure, we determined enrichment of genes in known canonical pathways that would be associated with environmental exposures/differences and/or potential patient co-morbidities. Z-scores indicated predicted activation (positive z-score, orange) or inhibition (negative z-score, blue) of the pathway based on the expression of the genes in the pathway, in the directionality of AAs. Black striped bars indicated pathways where no z-score/predication was indicated due to insufficient evidence in the IPA knowledgebase. The starred red line indicates a p value cut of 0.05 (-log(0.05) = ~1.3).

Upon investigation of the 1071 genes associated with SRR, unsupervised hierarchical clustering revealed that cases were grouped by African ancestry, where we saw Ethiopian cases cluster distinctly from Ghanaian and AA cases (**Figure 6B**), and this is likely due to the significant fraction of shared genes from the two analysis approaches. However, after unsupervised clustering of the 751 genes unique to the SRR analysis, we found that the clustering pattern of our cases drastically changed (**Figure 6C**). Instead, we found a distinct cluster of AA cases, and a second cluster containing all African cases. The African cluster did separate into two subnodes, but were distinct from AA cases, largely driven by an upregulated gene signature found among AA and not seen among our African patients.

We hypothesized that this signature may be driven by environmental influences or co-morbidity status differences between AA and African cases, that were being detected at the tumor level. We identified several known canonical pathways that had significant enrichment among AAs compared to Ghanaians and Ethiopians (*p* < 0.05). Strikingly, a significant number of canonical pathways were related to comorbidities and environmental influences, which may reflect patient comorbidity health status in our cohort. Specifically, pathways related to cardiac function, adiposity/obesity related pathways, viral pathways, diabetes, and insulin signaling pathways were found to be activated among AA patients (**Figure 6D**).

## DISCUSSION

This study is the first comparative RNAseq study of TNBC that utilized an African-enriched cohort of east and west Africans with African Americans to discern the influence of the complex genetic admixture on TNBC tumor biology related to racial disparities. Previous comparative studies with similar study designs have either only used self-reported race as a proxy to infer shared biological differences or used global AFR ancestry to categorize patients into groups based on arbitrary majority ancestry thresholds. However, those methods do not consider the range of diversity in genetic origin of both AFR and EUR ancestry due to the unique social history and resulting genetic admixture across the African diaspora^33^. Our findings support the emerging notion that inclusion of multi-ethnic patient groups into genomics disparities research can have a transformative impact on cancer research. The unique composition of the African diaspora in patient ancestry across our international ICSBCS cohort continues to allow us access to a broad range of admixture and unique genetic drivers of disparities.

Our ancestry estimations revealed a broad range and distribution of African ancestry among self-reported AAs, ranging from 52.73% to 99.99%. Variation in European admixture among AA patients is expected and has been shown to differ regionally through the US, where less European admixture was reported among AA individuals in the southeast compared to the northeast or pacific northwest^32,33^. In our present analysis, we have shown a range of African versus European admixture among AA patients. Among our Ethiopian patients, the surprisingly high level of European admixture was almost equal to African ancestry, with a significant proportion of South Asian admixture also shown. These non-African ancestral origins in Ethiopians have been previously reported, where a significant proportion of mtDNA haplotypes^29^ and Y chromosome haplotypes^30^ represent non-African origins, and more recent genetic analysis shows up to 50% of non-African ancestry among Ethiopians^31^. Our sub-continental ancestry estimates indicated that the shared west African origins between our Ghanaian and AA patients corresponds with MSL ancestry (medians: 24.1%, and 19.7%, respectively). However, while the predominant AFR ancestry of AA and Ghanaian patients is generally west African, the regional origin of African ancestry was distinct between these groups. Specifically, Ghanaians were primarily represented by YRI ancestry with less than 0.01% ESN ancestry, and AA were primarily represented by ESN, with only two of 66 AA reporting YRI ancestry over 30% (AA median YRI 0.00%). This is a relevant distinction, given that most studies that sought to identify AA-specific risk alleles utilized a YRI reference genome as the template for imputing genotypes. Our work indicates the YRI genetic background is less appropriate for AA patients than other AFR genome references and this could adversely impact the relevance and rigor of genetic risk studies that utilize a single AFR genome reference. Given the presence of distinct AFR subpopulation gene signatures, our findings support the hypothesis that AFR subpopulations harbor population-specific genetic drivers of gene regulation that are relevant to disease pathology and therefore also disease risk.

In this study, we also found that distinct gene expression signatures in TNBC are associated with superpopulation African ancestry, as well as regional/national African ancestry. Over 600 genes associated with African ancestry, which clustered among Ghanaian and AA patients compared to Ethiopians, were enriched for functions of increased immune cell trafficking and activation of migration signals. A combination of genomic and classical IHC methods across independent cohorts verified AFR-associated higher infiltration of tumor-associated leukocytes in the TME, supporting previous studies that indicate higher levels of inflammation and immune cell enrichment in African patients^16,23^. The distinction of immune gene expression signatures, shared among patients with substantial west African ancestry, is supported by observations in human evolution research of differential immune responses between European and African populations^44,45^ that may be related to clinical consequences of the African-specific Duffy-null blood group status^6,18^. Also, the immune signals that were identified in our superpopulation African-associated genes, were also specifically associated with MSL ancestry (**Figure 4C-E, Figure 5B, respectively**). Therefore, the shared MSL ancestral origin of our cohort is likely the region harboring a genetic factor of immunological responses we observe. Further studies are needed to untangle the actual alleles that may be MSL-specific and functionally involved in immune responses.

Interestingly, no overlapping genes signatures were found between YRI and ESN, the predominant African origin of Ghanaians and AA, respectively. Only YRI and GWD associated genes overlap among the west African population-associated genes. This was surprising given the YRI and ESN populations are geographically closer and presumably would have more similar genetic backgrounds leading to the largest overlap in gene signatures. The lack of any overlap suggests there is significant population-level divergence of genetic drivers that direct gene expression signatures even within African nations. However, the only source of ESN ancestry in our cohort is derived from AA patients, who also have unique environmental influences mediating genetic impact on gene signatures^36^, and so the lack of overlap may be due to these mediating factors.

Epidemiological studies show that an individual’s disease is influenced by the intersection of their genetics and their environment, where genetic ancestry, lifestyle, neighborhood, and diet all play significant roles. Whereas SRR is typically used to define or characterize these factors in comparisons between different ancestry groups, for SRR groups like AA, range of genetic admixture among this population alone warrants further steps to be taken to not generalize the influence of genetic ancestry, especially as we are continually moving in the direction of precision medicine in cancer. In our previous and present work, we have highlighted that by using estimated genetic ancestry as a continuous variable in LR models to define African ancestry-associated genes, a significant proportion of genes are distinct compared to using typical SRR-based approaches. It is important to note that SRR should not be ignored in this context either, where in the present analysis the SRR-associated gene signature highlighted canonical pathways that seem to be influenced by individual’s environment. These exciting findings begs for further evaluation of individuals with TNBC across the African diaspora, as we have established additional working relationships with African partners through ICSBCS across African nations and expanding our analysis of AA individuals from our various recruitment sites across the US to characterize the influence of African ancestry and environmental differences across individuals with varying admixture and lifestyle exposures.

## METHODS

### Ancestry patient cohort

#### ICSBCS patient cohort

The International Center for the Study of Breast Cancer Subtypes (ICSBCS) biorepository represents the efforts an international consortium of breast cancer clinicians and researchers with the goal to characterize breast cancer disease in diverse populations worldwide. We have prospectively recruited breast cancer patients since 2006, where formalin-fixed paraffin embedded (FFPE) tumor tissue has been collected. Institutional Review Board (IRB) approval for utilization of biorepository samples was obtained from participating sites in the United States (Weill Cornell Medical College, New York City, NY; Henry Ford Health System, Detroit, MI; and University of Michigan, Ann Arbor, MI) and our international African partnering institutions (Komfo Anokye Teaching Hospital, Kumasi, Ghana and the Millennium Medical College St. Paul’s Hospital, Addis Ababa, Ethiopia). In the present study, TNBC tumor tissue was obtained from a total of 45 patients, including 9 AA, 3 EA, 12 Ghanaians and 21 Ethiopians (**Supplemental Figure S1**). Confirmation of TNBC diagnosis by IHC was completed for Ghanaian and Ethiopian cases at our ICSBCS US site locations in Michigan (University of Michigan, Henry Ford Health System) and New York (Weill Cornell Medical College)

#### UAB patient cohort

The UAB TNBC has been previously described^26^, and consists of a convenience cohort of retrospective FFPE TNBC tissue collected between 2000 and 2012 at the University of Alabama at Birmingham (UAB). Samples were collected and used under the UAB IRB. In the present study, samples were analyzed from 74 patients, including 42 AA and 32 EA patients (**Supplemental Figure S1**).

#### EIPM patient cohort

All samples were collected and used under the Weill Cornell Medical College IRB. In the present study, we have estimated ancestry from TNBC tissue of 13 patients, including 1 AA, 6 EA, 2 Asian and 4 patients who responded “other” or declined to provide race/ethnicity information (**Supplemental Figure S1**).

### RNA extraction from archival FFPE tissue

RNA was extracted from archival FFPE tissue using a modified QIAGEN Rneasy® FFPE kit protocol. Briefly, prior to deparaffinization of the FFPE tissue, the samples are incubated with 1X acidic antigen retrieval solution at 90C for 5 minutes. Following incubation, samples are cooled to room temperature, and any excess paraffin is removed from the tube. We then proceeded through the standard kit protocol. RNA yield was quantified using the Qubit® RNA Broad Range kit and Qubit® 4.0 fluorometer.

### RNA library preparation and sequencing

The quality of each RNA is assessed using RNA High Sensitivity Screen on TapeStation (Agilent Technologies). For RNA sequencing, 100 ng of total RNA molecules were used to construct libraries using Illumina TruSeq RNA Exome Library Prep Kit, following manufacturer’s protocols. The final libraries were then quantified using Agilent D1000 Screen Tape as well as sequenced on Illumina MiSeq V2 Micro Kit to assess insert sizes and integrity before sequencing on a high-throughput sequencer. Each library was normalized to 4 nM and pooled and sequenced on Illumina NextSeq500 High Output Kit (Illumina, San Diego, CA). All sequencing reads were converted to industry standard FASTQ files using BCL2FASTQ (version 1.8.4).

### RNAseq data processing and QC of samples

Raw RNAseq reads were assessed with Fast QC^46^ (version 0.11.8), and Trimmomatic^47^ (version 0.36) was utilized for read trimming and adapter removal. Reads were aligned using HISAT2^48^ (version 2.0.4) with the GrCh37 reference genome. Picard tools (version 2.18.3, https://broadinstitute.github.io/picard/) was used to pull alignment metrics for the samples, where a number of sequenced reads were found to have high levels of read duplication. Duplicate reads were removed using Picard, and only samples that had 10M reads post de-duplication were utilized in subsequent gene expression analyses.

### Ancestry estimation using variants called from RNAseq alignments

Ancestry proportion is determined by the software ADMIXTURE v1.3.0^49^, which uses a maximum likelihood-based method to estimate the proportion of reference population ancestries in a sample. We genotyped the reference markers generated from 1,964 unrelated 1000 Genomes project^28^ samples directly on the RNASeq samples using GATK pileup. Individuals from populations MXL (Mexican Ancestry from Los Angeles USA), ACB (African Caribbean in Barbados), and ASW (African Ancestry in Southwest US) were excluded from the reference due to being putatively admixed. The reference was further filtered by using only SNP markers with a minimum minor allele frequency (MAF) of 0.01 overall and 0.05 in at least one 1000 genomes superpopulation. Variants are additionally linkage disequilibrium (LD) pruned using PLINK v1.9^50^ with a window size of 500kb, a step size of 250kb and r^2^ threshold of 0.2, resulting in 122377 markers remaining. The analysis results in a proportional breakdown of each sample into 5 superpopulations (AFR, AMR, EAS, EUR, SAS) and 23 subpopulations (**Supplemental Table S1**).

### Gene expression quantification and differential gene analysis

Stringtie^48^ (version 1.3.3) was used to quantify gene expression from our de-duplicated aligned reads. Quantified genetic ancestry and self-reported race (SRR) groups were used to identify ancestry- or SRR-associated genes in our cohort, using logistic regression analysis comparing gene expression either with the continuous ancestry variable, or categorical SRR variable. Genes with a *p* < 0.01 were included in further analyses. Unsupervised hierarchical clustering of our gene lists were completed using JMP® Pro 16 (SAS Institute Inc., Cary, NC).

### Network analyses of differentially expressed genes

Ingenuity Pathway analysis software (Qiagen, version 01-16) was used to determine involvement of our gene lists in various canonical pathways, determine upstream regulators, and to draw *de novo* networks involving our gene lists. For each analysis and gene list, the log fold change was calculated based on the resulting node structure of the samples when the gene lists underwent unsupervised hierarchical clustering, as our ancestry associated versus SRR gene lists resulted in different clustering patterns of our samples.

### Tumor associated immune cell abundance in tumors using RNAseq deconvolution and enrichment methods

To determine estimated abundances to tumor-associated immune cell populations, we used the online CIBERSORTx^39^ platform (https://cibersortx.stanford.edu/) with our gene expression values as input. The LM22 signature matrix file was used as reference, and the estimation was completed with quantile normalization disabled (as recommended for RNAseq data) with 500 permutations. Only CIBERSORTx output with that was determined to be significant (*p* < 0.05) was included in our analyses.

We have additionally used xCell for deconvolution of immune and other cell populations from our bulk RNAseq data^40^. Normalized TPM expression was used as input for the xCell algorithm.

### Immunohistochemistry (IHC) of CD3 and FOXP3

Formalin-fixed paraffin embedded (FFPE) tumor block were obtained from the ICSBCS biorepository. Slide preparations were conducted through Henry Ford Health System Histology Core, using standard operating protocols. From the FFPE blocks, 4um sections were obtained. Multiplex staining was done using FOXP3 at 1:100 dilution (BioLegend Cat No. 320101) with CD3 pre-dilute (Agilent, IR503) as the antibody diluent.

### Tumor infiltrating leukocyte (TIL) analysis from IHC

TIL markers from multiplex IHC staining were analyzed using HALO software (V3, Indica Labs). Stained slides were electronically scanned using the Leica Aperio scanner and transferred into the HALO program. Positively stained tumor cells were annotated from hematoxylin and eosin staining and matched to a serial section with FOXP3 and CD3 multiplex staining. A custom algorithm optimized to detect color differences between the two markers was used to determine the number of positively stained cells for each marker. Positive tumor cells for each marker were divided by the total number of tumor cells and converted to a percent for subsequent data analysis.

### TNBC subtyping

To determine TNBC subtypes of our samples, we input gene expression values into the Vanderbilt TNBC type online tool (https://cbc.app.vumc.org/tnbc/)^42^. The TNBC subtypes IM and MSL have been determined to primarily represent infiltrating immune cells and tumor associated stroma, respectively, and therefore these calls are reassigned to their second most correlated call and significant call^43^. Unsure calls (UNS) are where multiple correlations are significantly associated with a tumor gene expression profile, and in our cohort, these were able to be resolved after disregarding IM and MSL calls.

As a supplementary validation method to the gene expression correlation-based Vanderbilt TNBC classification tool, a summarized ranks measure was computed using the original TNBC Subtypes signatures for all samples using normalized RNA-Seq expression data. TNBC Subtype signatures were obtained from Lehmann et al^42^. Across all samples, all genes expressed were ranked from low to high expression using the rank function in R statistical software with minimum rank method used to resolve duplicate expression ties. For each sample, ranks for each gene in the given subtype signature were extracted and a representative median of ranks for the gene signature was calculated to estimate the overall regulation of the signature with respect to the total expression. The TNBC subtype signature with max median signature rank per sample was the assigned TNBC subtype for the sample.

## Data Availability

The data generated in this study are not publicly available due to the nondisclosure of genetic data issued through the Ministries of Health at corresponding African sites. These data must be approved for use by these entities, but may be available upon reasonable request from the corresponding author.

## ACKNOWLEDGMENTS

We would like to thank all the members of the International Center for the Study of Breast Cancer Subtypes, from the US and Africa, for their dedication to our mission. We would also like to extend our most sincere gratitude to all the patients and their families for their contribution and trust in this work.

