## Supplemental Figures for "African-Ancestry Associated Gene Expression Signatures and Pathways in Triple Negative Breast Cancer, a Comparison across Women of African Descent"

[1]Department of Surgery, Weill Cornell Medical College, New York City, NY, USA; (RM), (BL), (KG), (LN), (MD)
[2]Department of Genetics, University of Georgia, Athens, GA, USA
[3]Englander Institute for Precision Medicine, Weill Cornell Medical College, New York City, NY, USA; (PD), (AV), (OE)
[4]New York Genome Center, New York City, NY, USA; (TC), (KA), (NR)
[5]Department of Public Health Sciences, Henry Ford Health System, Detroit, MI, USA; (YC), (LJ), (EJ)
[6]Department of Biochemistry, St Paul's Hospital Millennium Medical College, Addis Ababa, Ethiopia; (EG)
[7]Department of Surgery, Komfo Anokye Teaching Hospital, Kumasi, Ghana; (JO), (FA)
[8]Department of Pathology, Komfo Anokye Teaching Hospital, Kumasi, Ghana; (EA),
[9]Department of Pathology, St. Paul's Hospital Millennium Medical College, Addis Ababa, Ethiopia; (AJ)
[10]Directorate of Oncology, Komfo Anokye Teaching Hospital, Kumasi, Ghana; (BA), (EO)
[11]Department of Surgery, St. Paul's Hospital Millennium Medical College, Addis Ababa, Ethiopia; (MB), (EA)
[12]Department of Surgery, Kwame Nkrumah University of Science and Technology, Kumasi, Ghana; (IK), (MA)
[13]Directorate of Radiology, Komfo Anokye Teaching Hospital, Kumasi, Ghana; (KA)
[14]Department of Pathology, Henry Ford Health System, Detroit, MI, USA; (DC)
[15]Department of Surgery, Henry Ford Health System, Detroit, MI, USA; (JB), (SN), (LP), (EP)
[16]Department of Translational Genomics, Keck School of Medicine, University of Southern California, Los Angeles, CA, USA; (LG), (ZM), (JC)
[17]Department of Population Sciences, City of Hope, Duarte, CA, USA; (RK)
[18]Department of Biology, Tuskegee University, AL, USA; (JW)
[19]Center for Cancer Research, Tuskegee University, AL, USA; (CY)

### Supplemental Figures

Supplemental Figure S1

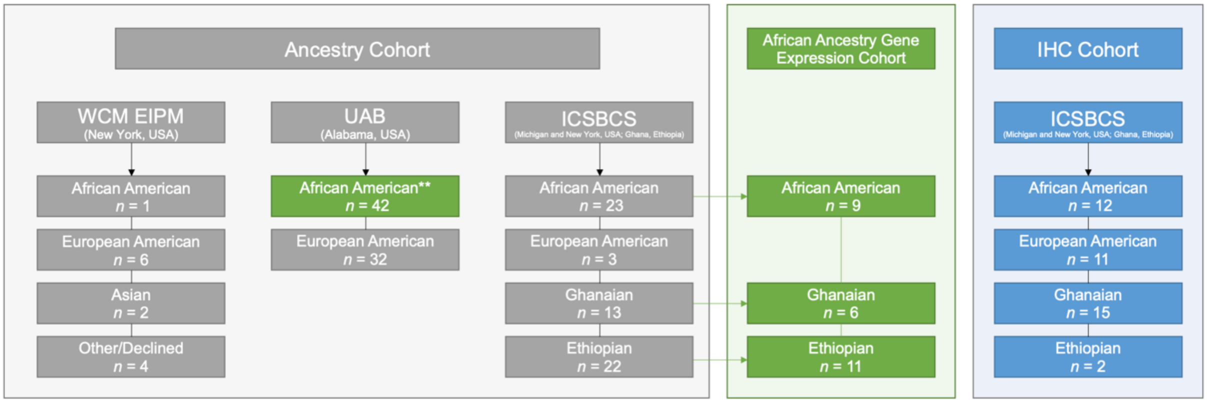


**Supplemental Figure S1. Ancestry and gene expression cohort composition.** Flow chart describing cohort and self-reported race breakdown of our ancestry cohort (gray) and gene expression cohort (green), and IHC cohort (blue) used for the present analysis. **Samples used for validation analysis of African subgroup associations.

Supplemental Figure S2

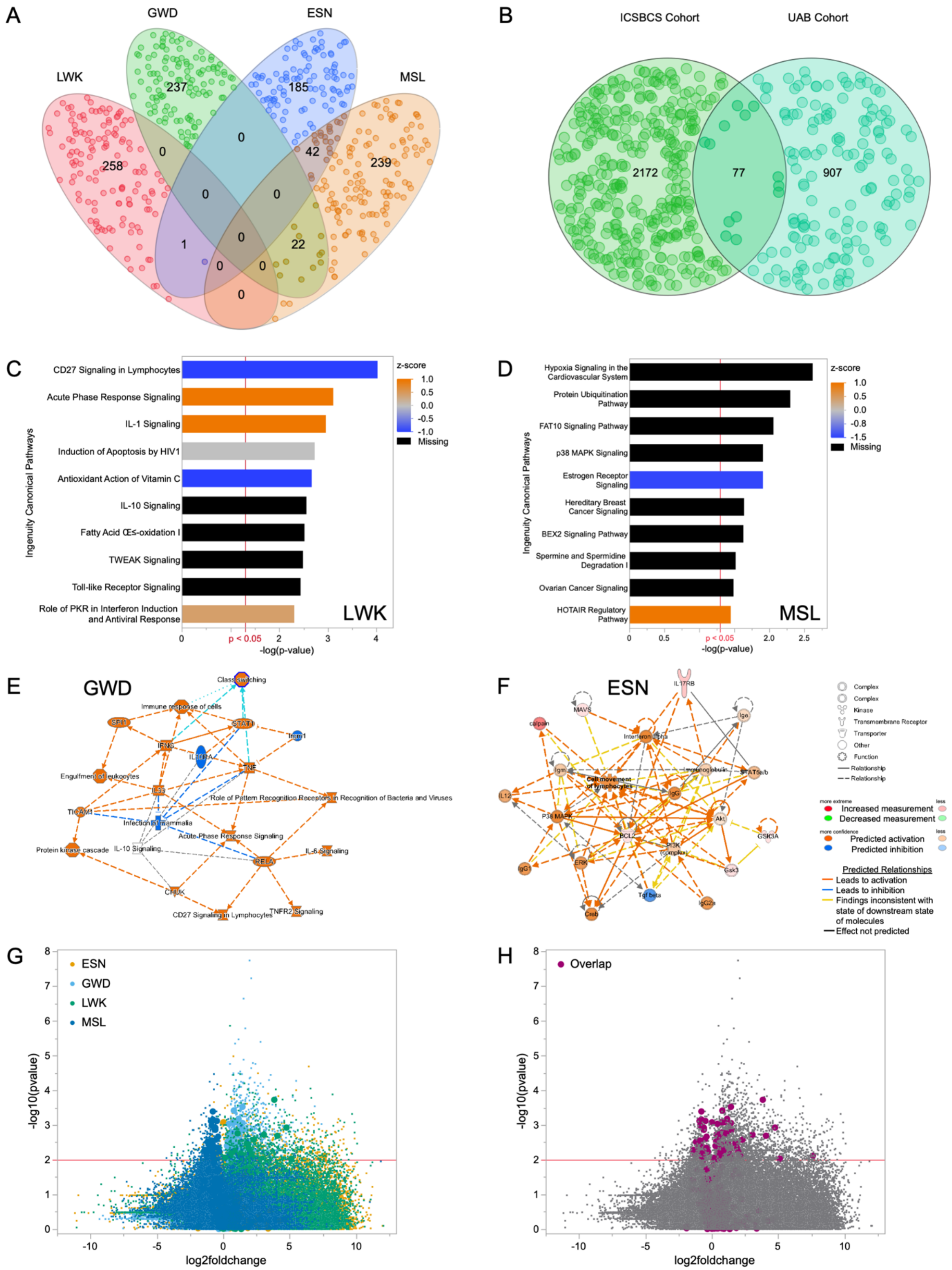

**Supplemental Figure 2. Replication of African subpopulation ancestry associated gene expression with AA patients in Davis and Martini et al[26].** (A) Venn Diagram showing overlapping genes in AFR subpopulation-associated gene signatures in the UAB cohort. (B) Venn Diagram showing overlapping genes between AFR subpopulation-associated gene signatures from the ICSBCS and the UAB cohort. Canonical pathway enrichment bar charts of IPA output from (C) LWK-associated gene signature and (D) MSL-associated gene signature. The y-axis lists canonical pathways that were enriched, and the x-axis shows the significance of the enrichment. Z-score showing predicted activation (orange) or inhibition (blue) is also shown. Bars in black have no predicted activation or inhibition due to insufficient evidence in the IPA knowledgebase. The starred red line indicates a p value cut of 0.05 (-log(0.05) = ~1.3). (E) Graphical summary of GWD-associated genes, and (F) network of subset of ESN-associated genes. Molecules in red and green are up- or downregulated, respectively, and those predicted to be activated (orange) or inhibited (blue) are also shown. (G) and (H) are volcano plot of AFR subpopulation-associated genes, where in (G) genes are color coded by AFR subpopulation (ESN = yellow, GSN = light blue, LWK = green, MSL = dark blue), and in (H) overlapping genes from (B) are highlighted in purple.

**Supplemental Figure S3. Correlation of xCell cellular populations and African ancestry.** Correlation of African ancestry and xCell[40] deconvoluted populations. Significant correlations are highlighted in shades of red.

Supplemental Figure S3

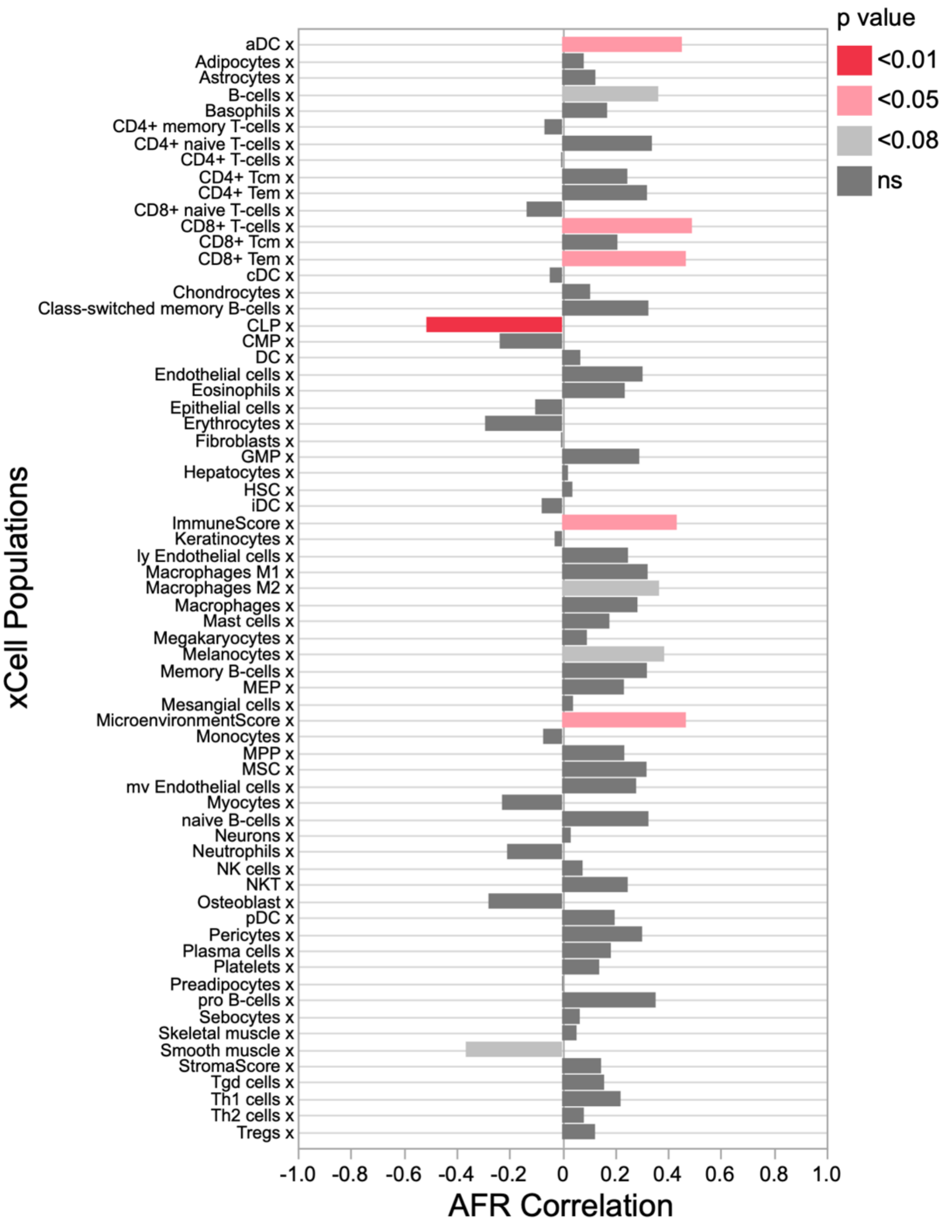

**Supplemental Figure S4. Population-level comparison of BMI reported across the ICSBCS cohort.** (A) Boxplots of BMI across SRR groups. N of each population reported. BMI of 30+ denoted obese individuals, 25-30 are overweight, 18.5-24.99 are normal BMI, and <18.5 are underweight. (B) P values of paired student's t-test comparing BMI between the indicated SRR groups.

Supplemental Figure S4

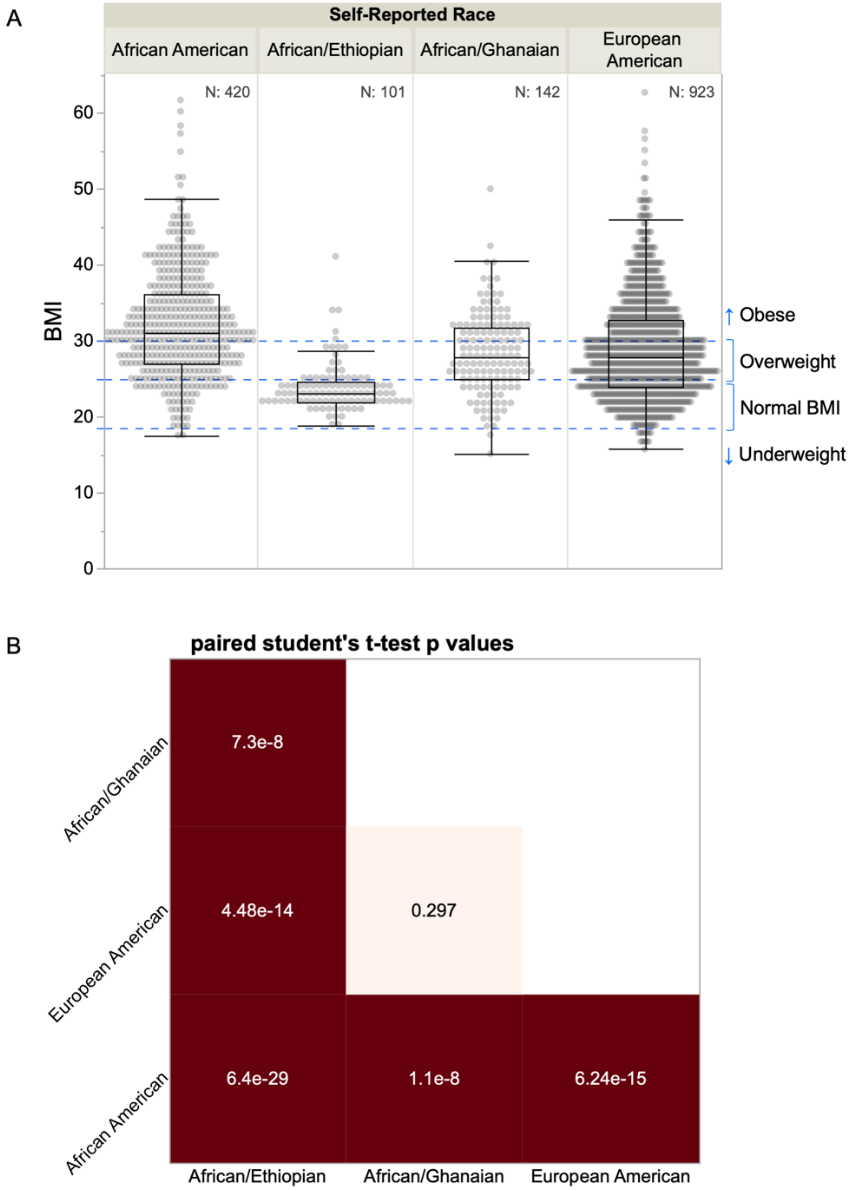
