## Supplemental Tables 1-2 for "African-Ancestry Associated Gene Expression Signatures and Pathways in Triple Negative Breast Cancer, a Comparison across Women of African Descent"

**Supplemental Table S1. 1000 Genomes Populations used as Reference for Ancestry Estimations**

| Superpopulation Population | Population Code | Population |
|---|---|---|
| East Asian (EAS) | CDX | Chinese Dai in Xishuangbanna, China |
| | CHB | Han Chinese in Beijing, China |
| | CHS | Southern Han Chinese |
| | JPT | Japanese in Tokyo, Japan |
| | KHV | Kinh in Ho Chi Minh City, Vietnam |
| South Asian (SAS) | BEB | Bengali from Bangladesh |
| | GIH | Gujarati Indian from Houston, Texas |
| | ITU | Indian Telugu from the UK |
| | PJL | Punjabi from Lahore, Pakistan |
| | STU | Sri Lankan Tamil from the UK |
| European (EUR) | CEU | Utah Residents (CEPH) with Northern and Western European Ancestry |
| | FIN | Finnish in Finland |
| | GBR | British in England and Scotland |
| | IBS | Iberian Population in Spain |
| | TSI | Toscani in Italia |
| American (AMR) | CLM | Colombians from Medellin, Columbia |
| | PEL | Peruvians from Lima, Peru |
| | PUR | Puerto Ricans from Puerto Rico |
| African (AFR) | ESN | Esan in Nigeria |
| | GWD | Gambian in Western Divisions in the Gambia |
| | LWK | Luhya in Webuye, Kenya |
| | MSL | Mende in Sierra Leone |
| | YRI | Yoruba in Ibadan, Nigeria |

**Supplemental Table S2: Median percent ancestry across 1000 Genomes superpopulations and subpopulations**

| | **African/Ghanaian**<br>*n* = 12<br>Median (min-max) | **African American (AA)**<br>*n* = 52<br>Median (min-max) | **African/Ethiopian**<br>*n* = 21<br>Median (min-max) | **European American (EA)**<br>*n* = 39<br>Median (min-max) |
|---|---|---|---|---|
| **East Asian (EAS)** | **1.25% (0.00% - 6.38%)** | **0.01% (0.00% - 18.78%)** | **0.00% (0.00% - 9.17%)** | **0.00% (0.00% - 40.74%)** |
| CDX | 0.00% (0.00% - 2.13%) | 0.00% (0.00% - 3.93%) | 0.00% (0.00% - 6.07%) | 0.00% (0.00% - 6.41%) |
| CHB | 0.00% (0.00% - 1.02%) | 0.00% (0.00% - 2.68%) | 0.00% (0.00% - 2.14%) | 0.00% (0.00% - 0.39%) |
| CHS | 0.00% (0.00% - 1.77%) | 0.00% (0.00% - 8.66%) | 0.00% (0.00% - 1.99%) | 0.00% (0.00% - 2.54%) |
| JPT | 0.00% (0.00% - 1.70%) | 0.00% (0.00% - 8.85%) | 0.00% (0.00% - 3.00%) | 0.00% (0.00% - 49.76%) |
| KHV | 0.00% (0.00% - 7.43%) | 0.00% (0.00% - 13.65%) | 0.00% (0.00% - 6.13%) | 0.00% (0.00% - 5.02%) |
| **South Asian (SAS)** | **0.64% (0.00% - 4.61%)** | **0.98% (0.00% - 16.72%)** | **8.99% (0.00% - 22.99%)** | **0.00% (0.00% - 14.27%)** |
| BEB | 0.00% (0.00% - 0.91%) | 0.00% (0.00% - 19.43%) | 0.00% (0.00% - 8.97%) | 0.00% (0.00% - 2.74%) |
| GIH | 0.00% (0.00% - 0.77%) | 0.00% (0.00% - 1.69%) | 0.00% (0.00% - 6.38%) | 0.00% (0.00% - 10.36%) |
| ITU | 0.00% (0.00% - 0.00%) | 0.00% (0.00% - 3.56%) | 0.00% (0.00% - 2.93%) | 0.00% (0.00% - 20.24%) |
| PJL | 0.00% (0.00% - 3.33%) | 0.00% (0.00% - 8.80%) | 0.00% (0.00% - 21.81%) | 0.00% (0.00% - 30.02%) |
| STU | 0.00% (0.00% - 0.00%) | 0.00% (0.00% - 6.52%) | 0.00% (0.00% - 15.82%) | 0.00% (0.00% - 6.77%) |
| **European (EUR)** | **0.00% (0.00% - 9.60%)** | **13.48% (0.00% - 51.10%)** | **43.52% (0.00% - 88.53%)** | **93.96% (8.12% - 99.99%)** |
| CEU | 0.00% (0.00% - 2.35%) | 0.00% (0.00% - 44.53%) | 0.00% (0.00% - 34.14%) | 9.65% (0.00% - 99.98%) |
| FIN | 0.00% (0.00% - 8.63%) | 0.00% (0.00% - 24.25%) | 0.00% (0.00% - 3.96%) | 0.00% (0.00% - 56.82%) |
| GBR | 0.00% (0.00% - 0.00%) | 0.00% (0.00% - 25.85%) | 0.00% (0.00% - 9.59%) | 0.00% (0.00% - 99.98%) |
| IBS | 0.00% (0.00% - 3.85%) | 0.00% (0.00% - 22.89%) | 0.00% (0.00% - 52.02%) | 0.00% (0.00% - 49.78%) |
| TSI | 0.00% (0.00% - 4.37%) | 0.00% (0.00% - 23.23%) | 41.69% (0.00% - 52.02%) | 0.00% (0.00% - 99.98%) |
| **American (AMR)** | **0.00% (0.00% - 2.37%)** | **1.21% (0.00% - 21.81%)** | **1.81% (0.00% - 14.76%)** | **0.00% (0.00% - 6.14%)** |
| CLM | 0.00% (0.00% - 0.00%) | 0.00% (0.00% - 29.90%) | 0.00% (0.00% - 3.60%) | 0.00% (0.00% - 12.46%) |
| PEL | 0.00% (0.00% - 2.89%) | 0.00% (0.00% - 6.48%) | 0.00% (0.00% - 3.96%) | 0.00% (0.00% - 3.43%) |
| PUR | 0.00% (0.00% - 0.00%) | 0.00% (0.00% - 21.61%) | 0.00% (0.00% - 0.00%) | 0.00% (0.00% - 0.00%) |
| **African (AFR)** | **97.34% (86.06% - 99.99%)** | **82.59% (16.64% - 99.99%)** | **42.98% (6.65% - 94.22%)** | **2.35% (0.00% - 83.96%)** |
| ESN | 0.00% (0.00% - 8.51%) | 36.14% (0.00% - 87.84%) | 0.00% (0.00% - 9.73%) | 0.00% (0.00% - 55.59%) |
| GWD | 0.00% (0.00% - 33.25%) | 0.00% (0.00% - 41.88%) | 0.00% (0.00% - 34.71%) | 0.00% (0.00% - 2.87%) |
| MSL | 24.10% (0.00% - 96.95%) | 19.70% (0.00% - 90.1%) | 0.00% (0.00% - 61.23%) | 0.00% (0.00% - 80.72%) |
| YRI | 65.96% (0.00% - 95.51%) | 0.00% (0.00% - 80.50%) | 0.00% (0.00% - 10.99%) | 0.00% (0.00% - 29.18%) |
| LWK | 0.00% (0.00% - 20.52%) | 7.53% (0.00% - 62.40%) | 43.00% (3.03% - 51.14%) | 0.00% (0.00% - 24.63%) |
